# Evaluation of Myocardial Perfusion and Immune Cell Response in Cardiac Allograft Dysfunction of Heart-Transplant Patients

**DOI:** 10.1101/2020.01.28.20018168

**Authors:** Paul J. Kim, Francisco Contijoch, Gerald P. Morris, Darrin Wong, Neil C. Chi, Pourya Yarahmadi, Yuko Tada, Darren Salmi, Patricia Nguyen

**Affiliations:** Division of Cardiovascular Medicine, University of California, San Diego, San Diego, CA, United States; Department of Bioengineering, University of California, San Diego, San Diego, CA, United States; Department of Pathology, University of California, San Diego, San Diego, CA, United States; Division of Cardiovascular Medicine, Stanford University, Stanford, CA, United States; Department of Pathology, Stanford University, Stanford, CA

**Author notes:** Corresponding Author: Paul J. Kim, MD, 9452 Medical Center Drive, La Jolla, CA 92037-7411, pjk017 at health dot ucsd dot edu.

**Keywords:** cardiac allograft dysfunction, stress myocardial perfusion, cardiac allograft vasculopathy

## Abstract

**Background:** We investigated the myocardial perfusion differences and changes in immune cell response in heart-transplant patients with nonspecific graft dysfunction (NGD) compared to cardiac allograft vasculopathy (CAV) patients and normal heart-transplant patients.

**Methods and Results:** We prospectively studied 17 heart-transplant patients (59.8±14.1 years, 78% male) from January to June 2016. Regadenoson stress cardiac MRI was performed in the patients and peripheral blood obtained contemporaneously to isolate peripheral blood mononuclear cells (PBMCs). Stress myocardial perfusion showed significantly decreased myocardial perfusion using maximum upslope method in NGD and CAV patients compared to normal heart-transplant patients. Myocardial scar by late gadolinium enhancement also was significantly increased in nonspecific graft dysfunction patients compared to normal. Evaluation of PBMCs by flow cytometry showed a trend towards increased activated HLA-DR^+^ T cells in NGD patients compared to normal. Clinical outcomes for cardiac hospitalization, allograft loss/retransplant, death were assessed at 8 years.

**Conclusions:** NGD shows decreased stress myocardial perfusion by cardiac MRI and a trend towards increased activated T cells in PBMCs, suggestive of an immune-mediated cause for allograft dysfunction.

## BACKGROUND

Despite improved recognition and understanding of rejection, a significant percentage of heart-transplant patients with graft dysfunction still do not have an identified cause. The major known causes of graft dysfunction beyond the first 24 hours post-heart transplant are acute cellular rejection (ACR), antibody-mediated rejection (AMR) and macrovascular cardiac allograft vasculopathy (CAV) ^1^. Heart-transplant patients are at highest risk of death from either ACR or AMR typically within the first three years after transplantation ^1^. Macrovascular CAV is seen later, often diagnosed years 5-10 after transplantation, and is widely recognized as the major limitation to long-term success of heart transplantation, limiting median longevity to 10 years ^1 , 2^. However, up to 36% of heart-transplant patients with graft dysfunction do not have an identified cause and are categorized as non-specific graft dysfunction (NGD) ^3^. These patients are typically diagnosed from years 1-10 after heart transplantation and actually represent the majority of morbidity and mortality during this time, accounting for 20-25% of deaths ^1, 3^. Our inability to define the cause of nonspecific allograft dysfunction is due to the limitations of the current tools used to detect rejection, specifically endomyocardial biopsy and invasive coronary angiography (ICA) ^4, 5^. Intravascular ultrasound (IVUS), the current gold standard for detection of CAV, is also limited as it cannot assess involvement of distal vessels and microvasculature ^6^.

Cardiac MRI (CMR) provides a comprehensive evaluation of heart-transplant patients with assessment of myocardial perfusion reserve, cardiac structure and function at a high spatial resolution, and myocardial edema and fibrosis. Microvascular disease in heart-transplant patients determined by invasive physiologic assessment (e.g., index of microcirculatory resistance) is associated with increased mortality and subsequent development of macrovascular CAV ^6–8^. However, assessment for microvascular disease is not commonly performed because of increased time required by the procedure and expertise is limited to only a few centers in the US. Myocardial perfusion reserve using stress perfusion CMR strongly correlates with the index of microcirculatory resistance and provides an alternative non-invasive methodology for detection of microvascular CAV ^5^.

Regadenoson is a newer selective A_2A_ adenosine receptor agonist used for vasodilator stress testing. Studies in non-transplant patients showed a significantly decreased incidence of high-degree atrioventricular block with regadenoson compared to adenosine infusion and in fact, recent studies showed no incidence of high-degree atrioventricular block with regadenoson ^9, 10^. The safety in heart-transplant patients was further demonstrated by Cavalcante and colleagues with no episodes of bradycardia or atrioventricular block with regadenoson ^11^. In contrast, there was a 12% incidence of second-degree atrioventricular block and 8% incidence of a sinus pause with adenosine. More recently, Kazmirczak and colleagues demonstrated the safety of regadenoson stress CMR scans in heart-transplant patients with no differences in the rates of adverse effects between heart-transplant patients and non-heart transplant patients ^12^. Moreover, an abnormal regadenoson stress CMR was associated with a significantly higher incidence of the composite outcome that included percutaneous coronary interventions, cardiac hospitalizations, retransplantations and deaths.

There is a broad consensus that macrovascular CAV is primarily due to a persistent T-cell mediated immune response that is resistant to calcineurin-inhibitors ^13^. Activated CD4^+^ T cells are predominantly present in the neointima and adventitia of macrovascular CAV lesions ^14, 15^. Additionally, these populations of T cells have been found to be oligoclonal in hearts with severe macrovascular CAV ^16^. Allorecognition can occur as direct (display of peptides by nonself HLA molecules expressed by graft-derived APCs) or indirect recognition (display of allogeneic peptide antigens by host APCs) ^13^. Thus, in this study, we looked to determine whether a T-cell mediated immune response is responsible for NGD, similar to macrovascular CAV, and examine whether activated T-cells in peripheral blood mononuclear cells provide the specific T-cell mediated mechanism for NGD.

## METHODS

### Patients

Adult heart-transplant patients at Stanford University Medical Center, Stanford, CA, USA and University of California San Diego, San Diego, CA, USA, were enrolled between January 2016 to December 2018. We identified heart-transplant patients who had nonspecific graft function (NGD), graft dysfunction due to macrovascular CAV or patients with normal graft function. Normal graft function is defined as having a left ventricular ejection fraction (LVEF) equal to or greater than 55% and no prior history of clinically significant acute rejection episodes that required modification of the immunosuppressive regimen or CAV. Patients with graft dysfunction due to macrovascular CAV are defined as having a LVEF equal to or less than 50% AND decrease from post-transplant baseline LVEF by an absolute difference of 10% or greater by echocardiography within a year of enrollment with diagnosis of macrovascular CAV by either ICA or IVUS as previously described ^17^. NGD patients are defined as having LVEF equal to or less than 50% AND decrease from post-transplant baseline LVEF by an absolute difference of 10% or greater by echocardiography within a year of enrollment, no diagnosis of macrovascular CAV ^17^, and no history of prior acute rejection episodes known to have decreased the LVEF to or less than 50%. Patients determined to have NGD previously underwent coronary angiography and IVUS in addition to endomyocardial biopsy. Patients enrolled did not have a contraindication to MRI, gadolinium contrast or regadenoson. Regadenoson stress CMR were performed at Stanford University Medical Center, Stanford, CA, USA and University of California (UC San Diego), San Diego, CA, USA. Additionally, we obtained two 8 mL vials of whole blood at the time of the intravenous catheter insertion for regadenoson stress CMR. Plasma and peripheral blood mononuclear cells (PBMC) were separated and stored from the whole blood samples. PBMCs were subsequently analyzed by flow cytometry to identify both activated CD4^+^ and CD8^+^ T cells. All CMR scans, collection, storage, and analysis of samples and patient data were performed under the approval of Stanford University and University of California San Diego human studies institutional review boards.

### Cardiovascular magnetic resonance

The CMR was performed using a 3T scanner (GE Signa Excite at Stanford University and GE 750 Discovery at UC San Diego) and a 32-element phased-array coil. All patients underwent a CMR protocol consisting of: 1) cine CMR at rest for assessment of left ventricular (LV) function; 2) gadolinium first pass perfusion imaging (basal, mid, and apical short-axis slices; every RR interval for 150 heart beats) 1 minute after regadenoson injection for assessment of stress perfusion; 3) gadolinium first-pass perfusion imaging (basal, mid, and apical short-axis slices; every RR interval for 150 heart beats) after reversal of regadenoson effect with intravenous caffeine for assessment of rest perfusion; 4) late gadolinium enhancement (LGE) CMR 10-15 minutes later; and 5) native and 20 minute post-gadolinium T1 map acquisition (mid short axis slice) for assessment of extracellular volume (ECV). The exam was completed in 45 minutes on average. Patients were instructed to avoid caffeine for 24 hours before the stress CMR study. Regadenoson 0.4 mg (Astellas, Northbrook, Illinois, USA) was injected over approximately 10 seconds into a peripheral vein followed by a 5 mL saline flush. Reversal of regadenoson was performed with 60 mg intravenous caffeine injected over 3 minutes ^18^. Gadolinium-based contrast (gadobenate dimeglumine, Bracco Imaging, Monroe Township, NJ) was infused at 2 ml/s using the dual bolus protocol previously described for both stress and rest perfusion ^19^. All patients were monitored by vector gating and pulse oximetry during the study. Blood pressure was monitored throughout the study with increased frequency at 3 minute interval for 10 minutes post regadenoson administration including reversal with intravenous caffeine. A 12-lead ECG was performed before and after the study.

### Image analysis

All stress CMR exams were analyzed blinded to patient outcomes by two readers (D.W. and P.K.) including LV mass, end-diastolic volume, end-systolic volume, LV and right ventricular (RV) ejection fraction, myocardial perfusion reserve by maximum slope, LGE mass by 6 standard deviation method and ECV by T1 mapping. Quantification of LV mass, end-diastolic volume, end-systolic volume, LV and RV ejection fraction, LGE mass and ECV were performed using cvi^42^ (Circle Cardiovascular Imaging, Calgary, AB, Canada).

### Perfusion quantification

Endocardial and epicardial contours were drawn on the perfusion images using cvi^42^. An additional region of interest was drawn in the blood pool, avoiding papillary muscles and trabeculae. The ROIs were manually translated on each perfusion image of the same slice to compensate for rigid-body translational motion. Perfusion quantification was performed in MatLab (MathWorks, Natick, Massachusetts) using algorithms written in house.

### Flow Cytometry

Human PBMCs were labeled with Live/Dead Yellow viability dye (Thermo-Fisher Scientific, Waltham, MA) before labeling with TruStain Fc block, anti-CD3 (HIT3a)-PE-Cy5, anti-CD4 (OKT4)-APC-Cy7, anti-CD8 (HIT8a)-PerCP-Cy5.5, and anti-HLA-DR-FITC (Biolegend, San Diego, CA). Samples were analyzed on a FACSCanto (BD Biosciences, San Jose, CA) with FACSDiva software. Samples were run in batches containing both control and experimental samples. Cutoffs for defining positive labeling were determined using fluorescence minus-1 controls for surface labeling. Data were analyzed using FlowJov10 (Tree Star, Ashland, OR).

### Assessment of clinical outcomes

Follow up data at 8 years after enrollment were collected through review of patient medical records at Stanford University and UC San Diego. Collected outcomes included: cardiac hospitalization, retransplantation and death. These events together formed the composite endpoint of major adverse cardiovascular events.

### Statistical analysis

All data were analyzed in a blinded fashion, with independent analysis of CMR and invasive data. Continuous variables are expressed as mean ± SD unless stated. Data were analyzed using Prism 6 software (GraphPad). Significant differences (P<0.05) were tested using ANOVA and Bonferroni post-test for > 2 groups. Nonparametric analyses were performed using Mann-Whitney test.

## RESULTS

### Patients

In total, seventeen heart-transplant patients were recruited. Of the seventeen patients, 8 had normal graft function (normal heart-transplant), 5 had NGD, and 4 had CAV (Table 1). There was no significant difference in age and gender between the groups. Regadenoson stress CMRs were performed earlier after heart-transplantation in normal heart-transplant and NGD patients while performed later for CAV patients (p = 0.001). More ACR episodes were seen in CAV patients (p = 0.03) and hyperlipidemia also more prevalent in CAV patients (p = 0.01). With regards to medications, ACE-inhibitors and beta blocker use was significantly increased in CAV patients compared to normal heart-transplant patients. Not surprisingly, mTOR inhibitor use was significantly increased in CAV patients compared to normal heart-transplant and NGD patients.

**Table 1.** Patient characteristics.

| Patient characteristic |  | Normal Heart-Transplant<br>(n = 8) | NGD<br>(n = 5) | CAV<br>(n = 4) | p value |
| --- | --- | --- | --- | --- | --- |
| Age, years (median, IQR) |  | 56 (52-63) | 60 (55-68) | 61.5 (53-70) | ns |
| Male, n (%) |  | 0.625 | 1 | 1 | ns |
| Graft age, years (median, IQR) |  | 2.125<br>(1.67-2.42) | 1.42<br>(1.08-7.42) | 10.42<br>(10.25-10.58) | 0.001 |
| ACR episodes, per patient |  | 0 | 0 | 0.5 | 0.03 |
| AMR episodes, per patient |  | 0 | 0 | 0 | ns |
| Hypertension, n (%) |  | 5 (62.5) | 4 (80) | 4 (100) | ns |
| Diabetes mellitus, n (%) |  | 3 (37.5) | 1 (20) | 3 (75) | ns |
| Hyperlipidemia, n (%) |  | 2 (25) | 2 (40) | 4 (75) | 0.01 |
| Serum creatinine, mg/dL (± SD) | | 1.32 (±0.4) | 1.82 (±0.66) | 1.09 (±0.24) | ns |
| Medications |  |  |  |  |  |
|  | Angiotensin converting enzyme-inhibitor, n (%) | 3 (37.5) | 4 (80) | 4 (100) | 0.04 |
|  | Angiotensin receptor blocker, n (%) | 0 (0) | 0 (0) | 0 (0) | ns |
|  | Beta-blocker, n (%) | 0 (0) | 0 (0) | 3 (75) | 0.001 |
|  | Calcium channel blocker, n (%) | 2 (25) | 1 (20) | 1 (25) | ns |
|  | Aspirin, n (%) | 8 (100) | 4 (80) | 4 (100) | ns |
|  | Statin, n (%) | 8 (100) | 5 (100) | 4 (100) | ns |
|  | Glucocorticoid, n (%) | 2 (25) | 1 (20) | 0 (0) | ns |
|  | Cyclosporine, n (%) | 0 (0) | 0 (0) | 1 (25) | ns |
|  | Tacrolimus, n (%) | 8 (100) | 4 (80) | 2 (50) | 0.027 |
|  | mTOR inhibitor, n (%) | 1 (12.5) | 2 (40) | 4 (100) | < 0.001 |

### Cardiovascular magnetic resonance

CMR data are presented in Table 2 and Figures 1 and 2. At the time of the CMR scan, NGD and CAV patients demonstrated significantly decreased LVEF compared to normal heart-transplant patients (Table 2). NGD patients also showed significantly decreased RVEF compared to normal heart-transplant patients. However, indexed LV volumes and mass showed no significant difference between the different groups.

**Figure 1.**
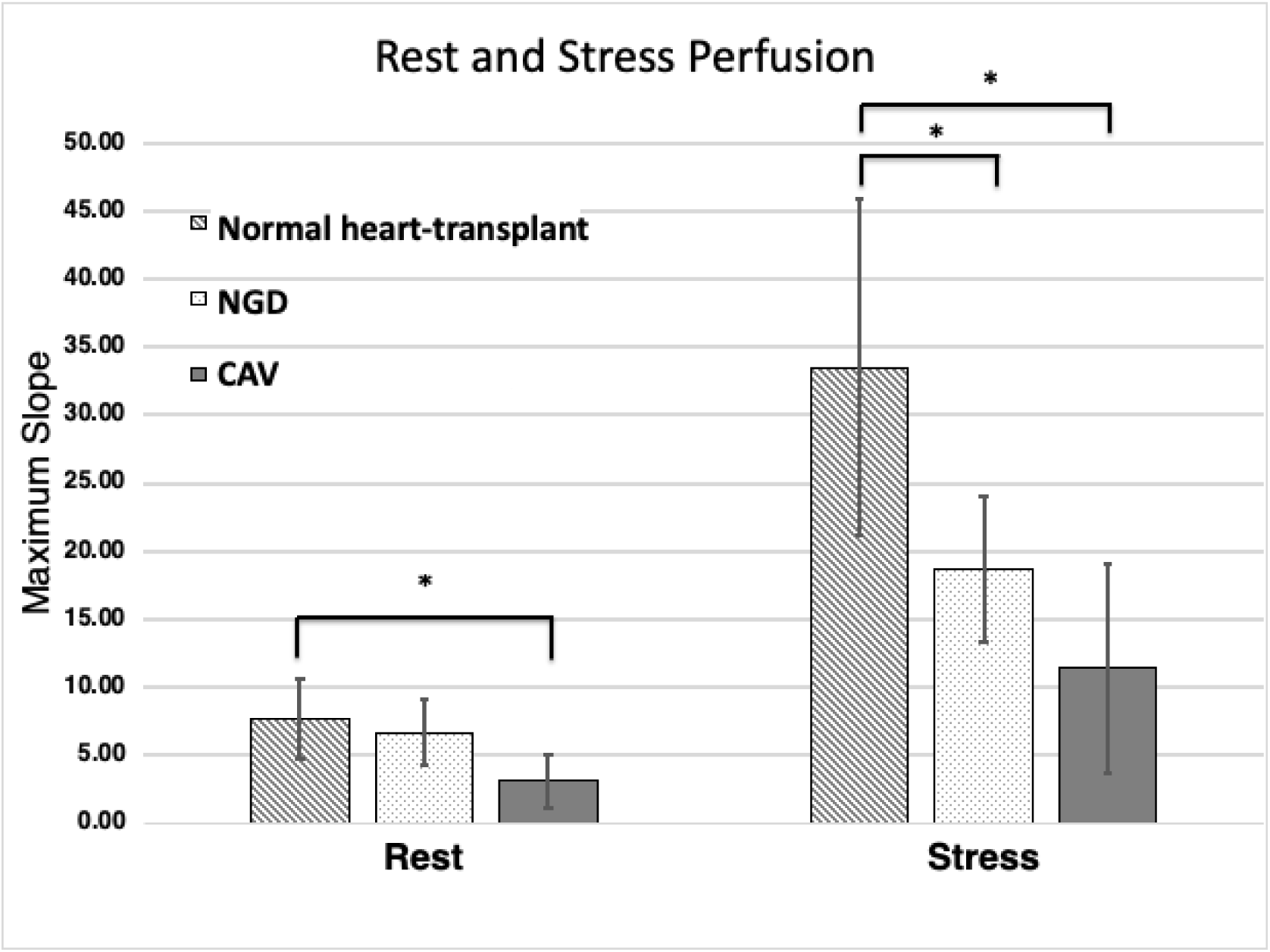
NGD patients demonstrated a trend toward decreased rest perfusion compared to normal heart-transplant patients while CAV patients showed a significantly decreased rest perfusion compared to normal. After stress with regadenoson, both NGD and CAV patients showed significantly decreased myocardial perfusion compared to patients with normal graft function. *, p < 0.05

**Figure 2.**
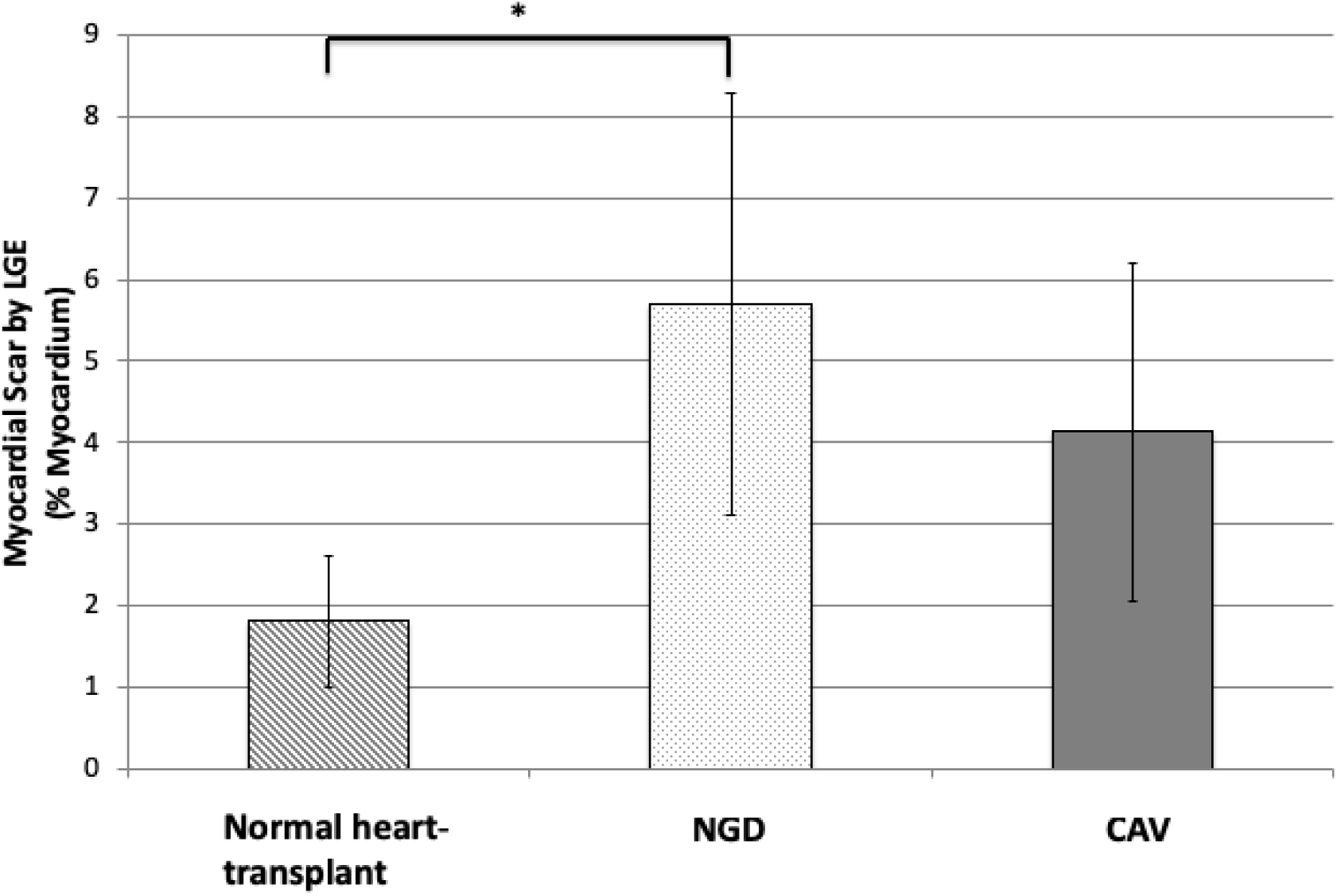
NGD patients showed significantly increased myocardial scar compared to normal heart-transplant patients. CAV patients also showed a trend towards increased myocardial scar compared to normal heart-transplant patients. * , p < 0.05

**Table 2.** Comparison of CMR parameters.

|  | Normal Heart-Transplant<br>(n = 8) | NGD<br>(n = 5) | CAV<br>(n = 4) | p value |
| --- | --- | --- | --- | --- |
| LVEF | 60.75 | 52.94 | 51.43 | 0.01 |
| RVEF | 49.88 | 42.56 | 55.58 | 0.03 |
| LVEDVI | 49.91 | 51.89 | 45.60 | ns |
| LVESVI | 19.69 | 24.14 | 21.92 | ns |
| LVMI | 51.16 | 56.12 | 56.99 | ns |

**Table 3.** Clinical outcomes.

|  | Normal Heart-Transplant<br>(n = 8) | NGD<br>(n = 5) | CAV<br>(n = 4) | *p* value |
| --- | --- | --- | --- | --- |
| Cardiac hospitalization | 0 | 0.2 | 1.5 | 0.04 |
| Graft loss/retransplant | 0 | 0 | 0 | ns |
| Death | 0.2 | 0.2 | 0.5 | 0.03 |
| Composite endpoint | 0 | 0.07 | 0.67 | 0.01 |

Resting myocardial perfusion is significantly decreased in CAV patients compared to normal heart-transplant patients (Figure 1). NGD patients showed a trend towards decreased resting myocardial perfusion compared to normal heart-transplant patients. After regadenoson, stress myocardial perfusion increased by a factor of six for normal heart-transplant patients. In contrast, stress myocardial perfusion is significantly decreased in both NGD and CAV patients compared to normal heart-transplant patients. There were no significant adverse effects with regadenoson administration in this study.

On evaluation of myocardial scar by LGE (Figure 2), NGD patients showed significantly increased myocardial scar compared to normal heart-transplant patients. While CAV patients also showed a trend towards increased myocardial scar, this result was not significantly different compared to normal heart-transplant patients. Figure 3 demonstrates an example of myocardial scar in the basal posterior wall visualized by LGE for an NGD patient with normal coronary arteries by ICA (Figure 4). In comparing myocardial edema by ECV (data not shown), no significant differences were seen between the different heart-transplant groups.

**Figure 3.**
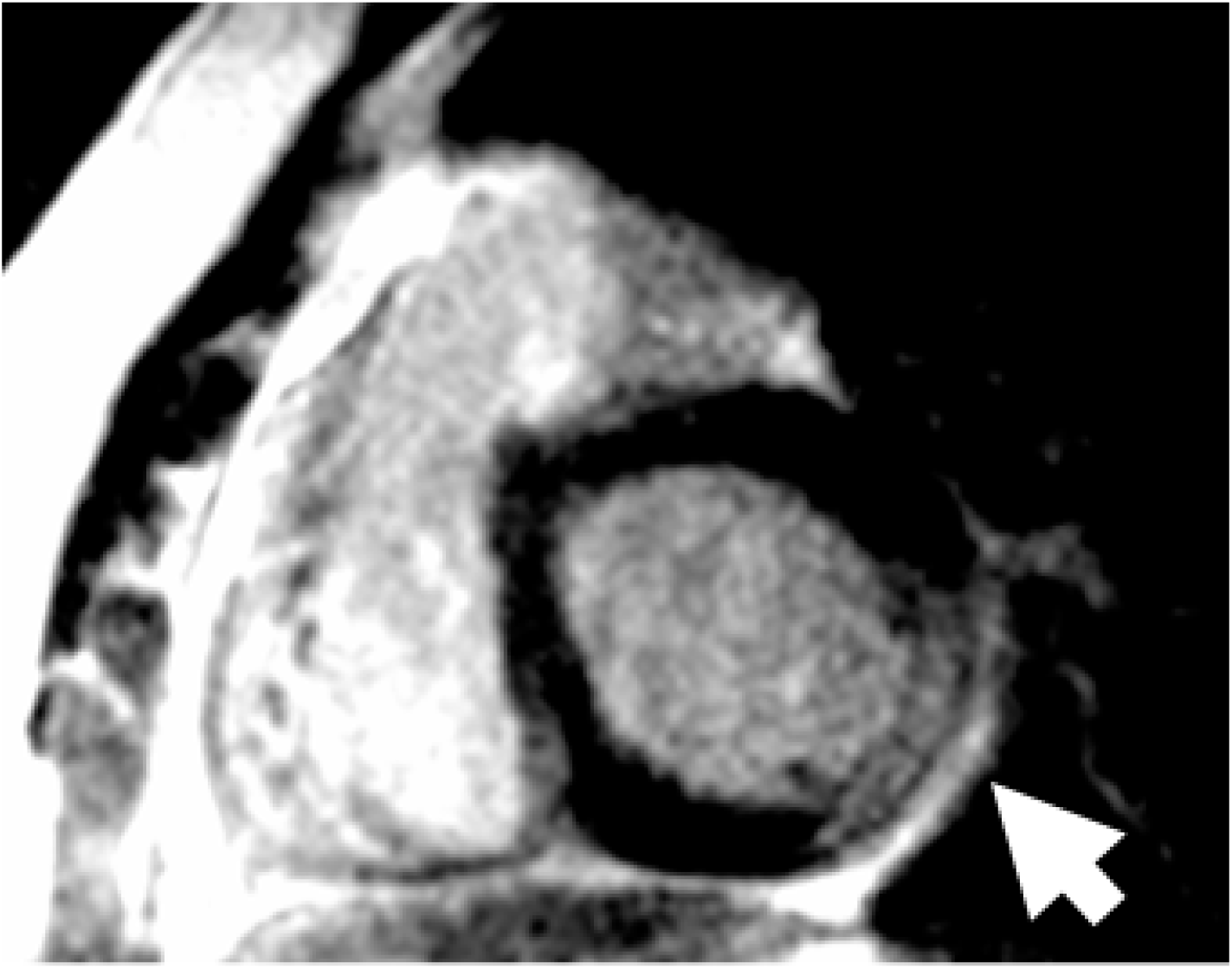
NGD patient demonstrates myocardial scar by LGE in the basal posterior wall (basal short axis view; RV=Right Ventricle, LV=Left Ventricle).

**Figure 4.**
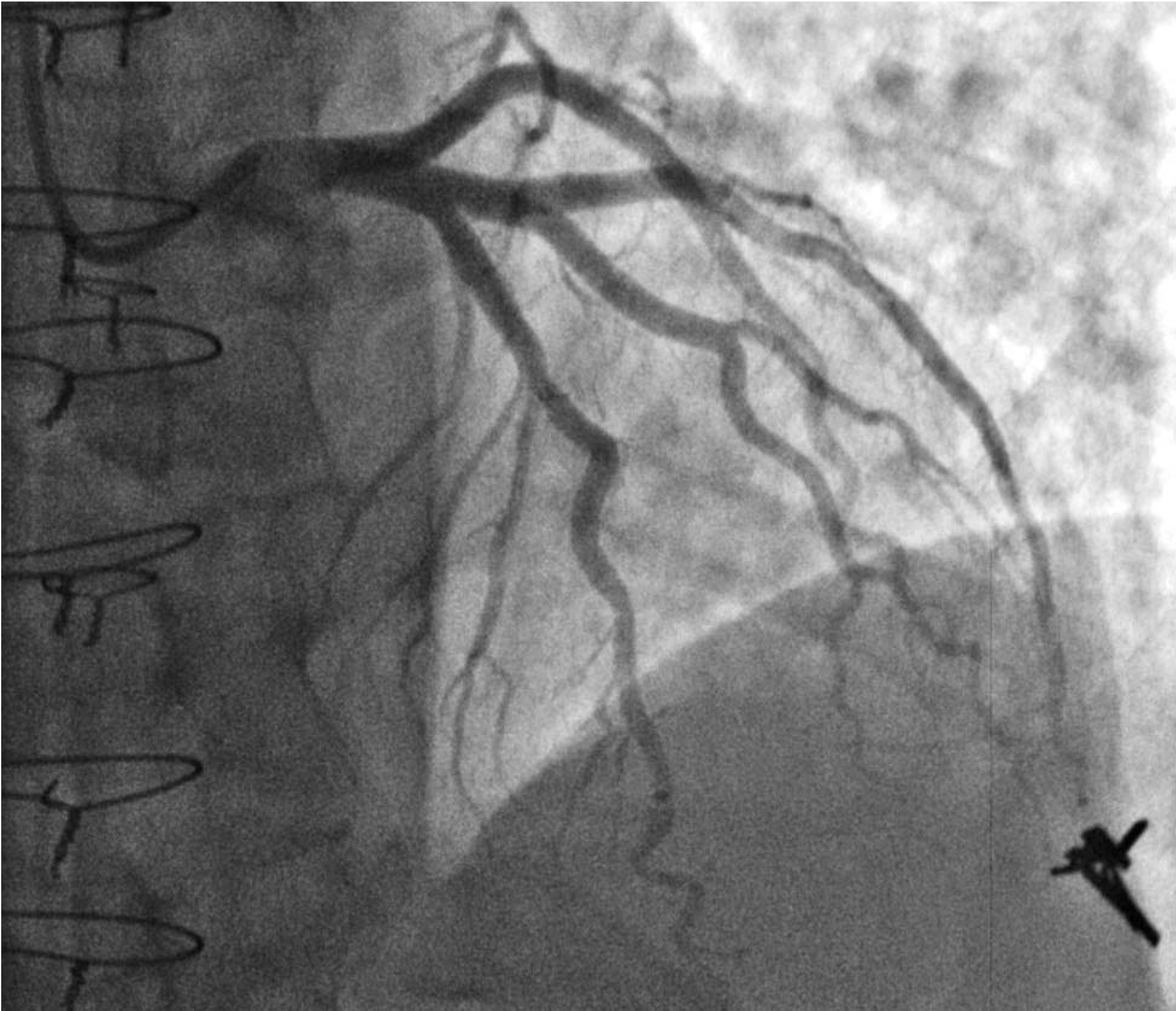
Invasive coronary angiogram in the same NGD patient. ICA demonstrates normal coronary arteries and IVUS (not shown) did not demonstrate intimal medial thickening to suggest early macrovascular CAV.

### Flow cytometry of PBMCs

After isolating T cells from PBMCs, we identified activated T cells by detection of HLA-DR^+^ using flow cytometry. We identified an important trend towards increased activated CD4^+^ and CD8^+^ T cells in NGD patients (Fig. 5 and 6). Figure 6 shows no difference of activated T cell populations in healthy control compared to normal heart-transplant patients, as expected. However, macrovascular CAV patients also do not show an increased activated T cell population compared to healthy control patients.

**Figure 5.**
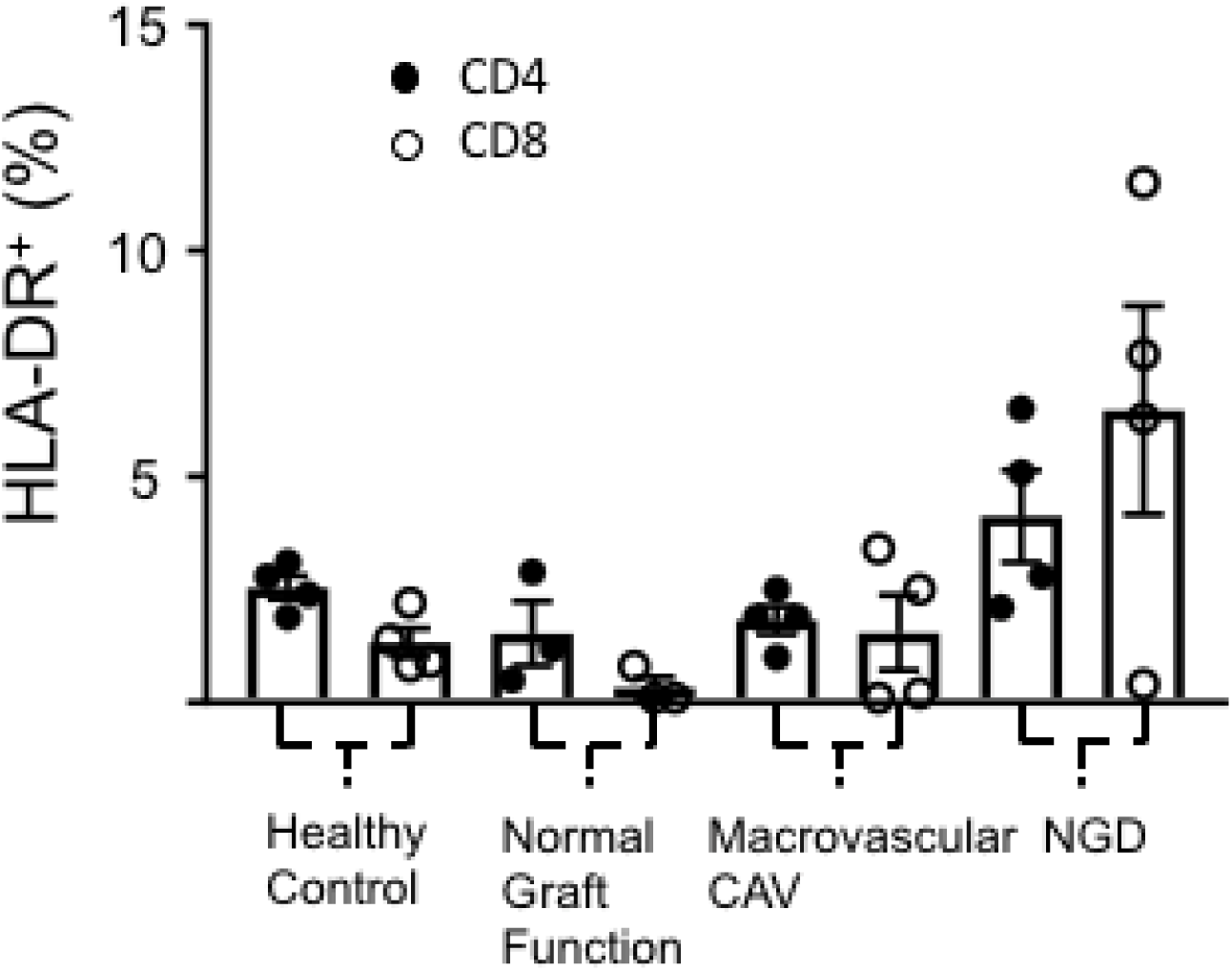
Trend showing increased percentage of activated CD4^+^ and CD8^+^ by flow cytometry in NGD patients compared to normal heart-transplant, CAV and healthy control patients. Activated T cells are indicated by detection of HLA-DR^+^. Data shown are the mean ± S.E.M.

**Figure 6.**
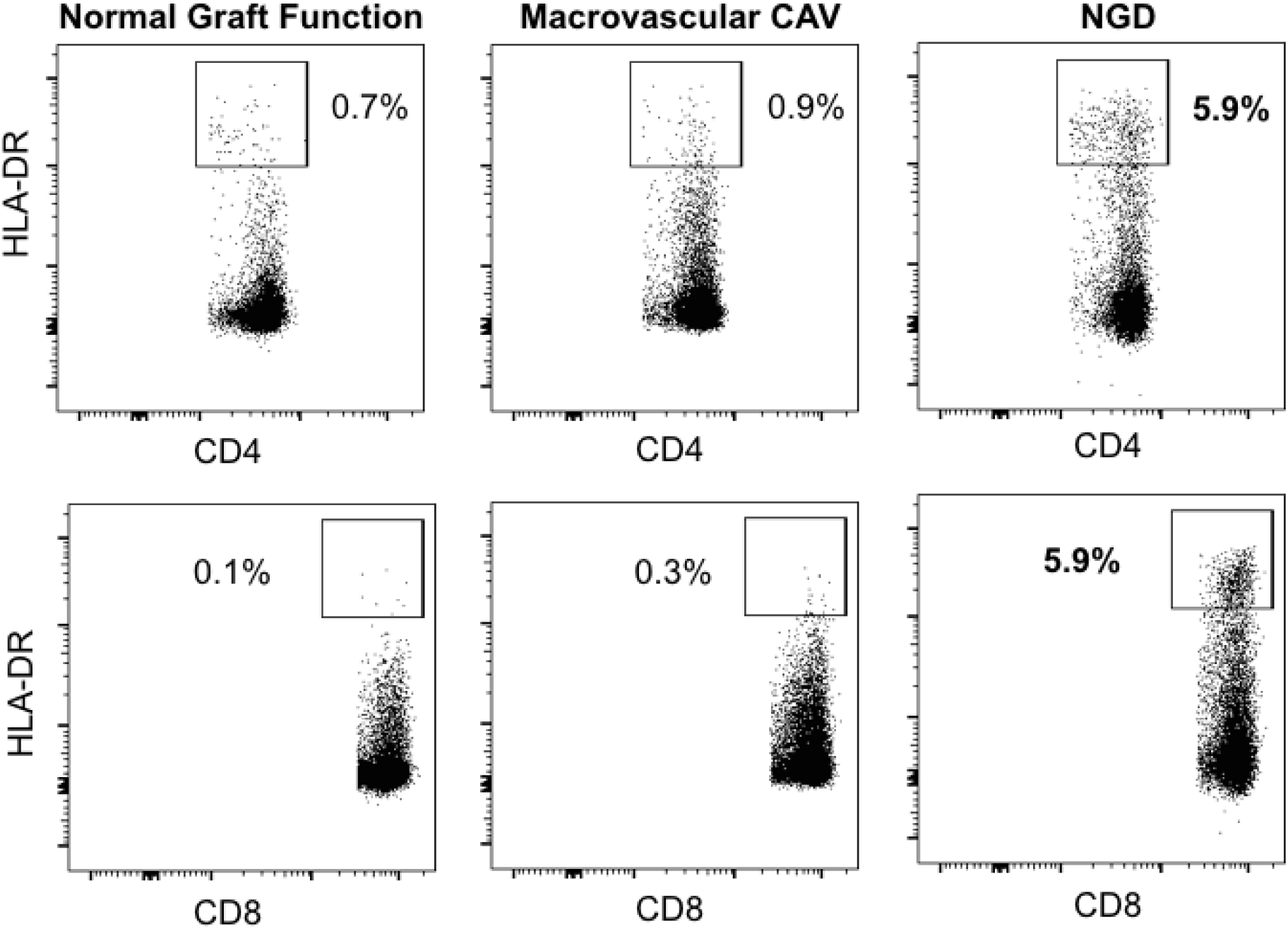
Representative PBMC samples by flow cytometry demonstrating increased percentage of activated CD4^+^ and CD8^+^ T cells in NGD patients compared to normal heart-transplant and CAV patients. Activated T cells indicated by detection of HLA-DR^+^.

### Clinical outcome

During a median follow-up of 7 years, there were no instances of graft loss/retransplantations, seven cardiac hospitalizations and four deaths. CAV patients demonstrated an average of 1.5 cardiac hospitalizations per patient with two patients accounting for six total hospitalizations. NGD patients had an average of 0.2 cardiac hospitalizations per patient with one cardiac hospitalization in the group. There were no hospitalizations for normal heart-transplant patients. There were two deaths in the CAV group, accounting for a mortality of 50% in the group. There was one death each in the NGD and normal heart-transplant groups.

## DISCUSSION

The principal finding of this study is that NGD patients demonstrate significantly decreased stress myocardial perfusion after regadenoson injection compared to normal heart-transplant patients. This is similar to that of decreased stress myocardial perfusion observed in CAV patients, although to a lesser degree. Interestingly, we see resting perfusion significantly decreased in CAV patients. In atherosclerotic coronary artery disease in non-transplant patients, myocardial perfusion is often “compensated” by an increased vasodilated state at rest, thus accounting decreased myocardial perfusion reserve as further vasodilation is limited with stress ^20^. However, we observe in heart-transplant patients with severe allograft vasculopathy that the resting myocardial perfusion is also significantly impacted and decreased. This abnormality may be explained by coronary endothelial dysfunction previously described in allograft vasculopathy ^7^. Though with stress, myocardial perfusion does increase in CAV patients, their stress myocardial perfusion is significantly decreased compared to normal heart-transplant patients as their resting myocardial perfusion is already profoundly depressed.

In NGD patients, there is a trend towards decreased resting perfusion. However, the decrease in stress myocardial perfusion truly differentiates these patients from normal heart-transplant patients. Thus, the stress state is needed to elicit lack of increased myocardial perfusion in response to increased metabolic demand in NGD patients. Also, as previously described, this decrease in myocardial perfusion is not correlated with increased myocardial scar/fibrosis, shown by LGE burden ^21^. As a result, the findings of this study point to decreased myocardial perfusion as one of the main culprits for graft dysfunction that we see in NGD patients, a group in which there is currently no attributed cause. As myocardial perfusion reserve by stress CMR has previously been shown to correlate highly with microvascular allograft vasculopathy in heart-transplant patients with normal epicardial coronary evaluation ^5^, results from this study suggest that microvascular allograft vasculopathy is the underlying pathologic cause for NGD. Significantly increased LGE burden, often in a coronary distribution, seen in NGD patients compared to normal heart-transplant patients provides further evidence to the hypothesis that microvascular allograft vasculopathy is the cause for NGD.

We further evaluated for a molecular basis for NGD by investigating the immune response between the groups. We observed that NGD patients showed a trend towards increased activated CD4^+^ and CD8^+^ T cells when compared to normal heart-transplant and CAV patients. We hypothesize that the ongoing inflammatory response by the activated CD4^+^ and CD8^+^ T cells accelerates microvascular disease, ultimately causing NGD due to microvascular graft vasculopathy. We suspect CAV patients do not demonstrate as much of an increase in activated CD4^+^ and CD8^+^ T cells due to the fact that the macrovascular graft vasculopathy has been ongoing so long (years) that both the allograft and the immune response has effectively “burned out.” CAV patients were scanned 9 years later than NGD patients after their heart-transplants, which is consistent with the usual onset of macrovascular CAV in literature and ISHLT registry data ^1^.

With respect to clinical outcomes, not surprisingly, we found that CAV patients showed the highest mortality and highest rate of cardiac hospitalizations. This is despite the fact that their graft dysfunction was similar to that of NGD patients. In comparison, NGD patients did have increased cardiac hospitalizations compared to normal heart-transplant patients but there was only one death in the NGD group. As previously described, CAV patients demonstrate structural and functional abnormalities much differently compared to non-transplant patients with coronary artery disease ^22^. In CAV patients, the LVEF can often remain preserved up until death or time of retransplantation and the LV does not usually dilate and remodel as we are accustomed to seeing in dilated ischemic cardiomyopathy in non-transplant patients. The ischemia from macrovascular CAV causes more of a restrictive cardiomyopathy with significant diastolic dysfunction in heart-transplant patients. This highlights the importance of earlier detection of myocardial ischemia in heart-transplant patients as LVEF is often not proportionately affected.

The clinical implications of these findings are that stress CMR using regadenoson potentially identifies microvascular allograft vasculopathy as the underlying pathologic cause of NGD patients. The importance of early detection of decreased stress myocardial perfusion is already shown in CAV patients, who are at high risk for sudden death despite relatively preserved LVEF. Finally, our study suggests an immune mediated mechanism for NGD through activated CD4^+^ and CD8^+^ T cells that may accelerate microvascular disease by chronic inflammation of the microvasculature.

Limitations of this study include its small sample size and the fact that this was a single center study. These findings need to be further validated in a larger study across multiple transplant centers. Further investigation of the molecular basis for NGD needs to be performed through more specific identification of the pathogenic T cells and elucidation of the pathway for immune-mediated injury.

## CONCLUSIONS

In NGD patients, myocardial perfusion at stress is significantly decreased compared to normal heart-transplant patients, consistent with an ischemic cause for graft dysfunction similar to that of CAV patients. Increased activated CD4^+^ and CD8^+^ T cells are shown in NGD patients that suggest an immune-mediated cause for vasculopathy and graft dysfunction.

## Data Availability

Our data is publicly available through Mendeley Data, an open access repository

## Acknowledgements.

We acknowledge Juan Santos, PhD, HeartVista, Los Altos, California, Reeve Ingle, PhD, Google, Mountain View, California and Burhan Jama, University of California San Diego, San Diego, California.

## Funding

The project described was partially supported by the National Institutes of Health (PJK), Grants UL1TR001442 and 1KL2TR001444, Astellas Pharma Investigator Sponsored Research, Stanford Translational Research and Applied Medicine Pilot Grant, and the American Heart Association Career Development Award 18CDA34110250. The content is solely the responsibility of the authors and does not necessarily represent the official views of the NIH.

## Authors’ contributions

GPM made substantial contributions to acquisition and interpretation of flow cytometry of PBMCs. DS made substantial contributions to the pathologic tissue analysis for microvascular changes. FC, DW, NC, PY, YT, PN, and PJK made substantial contributions to the interpretation of data for the work and revising it critically for important intellectual content. PJK made substantial contributions to the conception and design of the work; the acquisition, analysis and interpretation of the data for the work and to drafting the work and revising it critically for important intellectual content. All authors provided final approval of the version to be published.

## Ethics approval and consent to participate

The study was approved by Stanford University and University of California San Diego’s Institutional Review Board.

## Notes

### Competing Interest Statement

Paul Kim was supported by Astellas Pharma Investigator Sponsored Research. Astellas Pharma did not restrict review or publication of this data in any way.

### Author Declarations

All CMR scans, collection, storage, and analysis of samples and patient data were performed under the approval of Stanford University and University of California San Diego human studies institutional review boards.

### Summary of Updates

Updated co-authors and corrected title page

